# Detection of Monkeypox virus DNA in the wastewater of an airport in Rome, Italy: expanding environmental surveillance to emerging threats

**DOI:** 10.1101/2022.08.18.22278932

**Authors:** G La Rosa, P Mancini, C Veneri, G Bonanno Ferraro, L Lucentini, M Iaconelli, E. Suffredini

## Abstract

Since May 2022, monkeypox cases have been reported in non-endemic countries, and the disease was declared a Public Health Emergency of International Concern. Rapid identification of new cases is critical for outbreak control. Environmental surveillance can be used as a complementary tool for detecting pathogens circulating in communities. This study aimed to investigate whether Monkeypox DNA can be detected in wastewater from a transportation hub.

Twenty samples were collected in Italy’s first airport in Rome and tested using published and modified real-time PCR assays targeting the G2R region (TFN gene), F3L, and N3R genes. Nested PCR assays were also used for confirmation by sequencing. Three samples tested positive by real-time PCR and/or by nested PCR, confirming the occurrence of the virus in the airport’s wastewater.

Wastewater surveillance can be quickly adapted to investigate emerging threats and can be used to track the introduction and/or the diffusion of the Monkeypox virus in communities.

## Introduction

The monkeypox virus (MPXV), a member of the family Poxviridae, genus Orthopoxvirus, causes Monkeypox, a viral zoonosis, first detected in parts of northern Africa in the 1970s, closely related to smallpox, caused by the variola virus, eradicated in 1980 with subsequent cessation of vaccination. Animal hosts include different rodents and non-human primates. The virus is transmitted from one individual to another by close contact with lesions and by contact with body fluids, respiratory droplets, and contaminated materials such as bedding (WHO 2022a).

Monkeypox primarily occurs in central and west Africa, with periodic re-emergence of the disease. Since May 2022, an outbreak of monkeypox disease in non-endemic regions outside Africa has received considerable attention worldwide. Different WHO regions have been involved, mainly Europe and the list of countries affected by this poxvirus is growing. On 23 July 2022, Monkeypox was declared a Public Health Emergency of International Concern by the WHO (WHO 2022b; Ghebreyesus, 2022) after the H1N1 influenza pandemic (2009), poliovirus (2014), Ebola virus disease in west Africa (2014), Zika virus disease (2016), Ebola virus disease in the Democratic Republic of the Congo (2019), and COVID-19 in 2020 (Wenham et al., 2022). As of 9 August 2022, 17897 cases of Monkeypox have been recognized from 41 countries throughout the European region, according to the last ECDC-WHO report (ECDC-WHO, 2022). Of these, 17402 were laboratory confirmed, the earliest date of symptom onset reported as 3 April 2022. More than 500 cases have been reported in Belgium (546), Italy (599), Portugal (710), Netherlands (959), France (2423), United Kingdom (2973), Germany (2982), and Spain (5162). Rapid identification of new cases is critical for outbreak control. The environmental surveillance tool, which started with poliovirus monitoring and rose in popularity with Covid-19, has been recognized as a powerful tool for assessing pathogens circulation within the community. Following the EU Recommendation 2021/472, it has been successfully used to track SARS-CoV-2 and its variants across the EU countries. Infrastructures – both physical and virtual – required for wastewater-based disease surveillance have been created, and currently, about 1370 wastewater treatment plants are under systematic monitoring across the EU (European Commission, 2022).

Following the monkeypox outbreak that started in May 2022, some research groups involved in the environmental surveillance of SARS-CoV-2 expanded their efforts to investigate the virus’s occurrence in wastewater. Indeed, viral shedding DNA from saliva, feces, urine, semen, and skin lesions has been demonstrated (Peiró-Mestres et al., 2022; Lapa et al., 2022; Antinori et al., 2022), suggesting that Monkeypox genome is expected to be detected in wastewater. To date, no study in peer-reviewed journal articles has documented the occurrence of Monkeypox in sewage; however, two preprints have been published, one in the Netherlands and one in the United States, in western California (de Jonge et al., 2022; Wolfe et al., 2022).

In this study, we examined wastewater of the Fiumicino airport in Rome (the busiest airport in the country, the 8^th^ busiest airport in Europe) to investigate the occurrence of Monkeypox DNA as part of surveillance efforts to detect the spread of the virus. The usefulness of monitoring transportation hubs has already been demonstrated previously. In 2020, Ahmed and co-workers detected SARS-CoV-2 RNA in commercial passenger aircraft and cruise ship wastewater, suggesting wastewater surveillance from large transport vessels as a potential complementary data source to prioritize clinical testing and contact tracing among passengers (Ahmed et al. 2020). Later on, Agrawal and co-workers assessed the earliest introduction of the SARS-CoV-2 Omicron variant in Germany by monitoring a wastewater stream originating from Frankfurt airport using sequencing analysis (Agrawal et al., 2022). The European Centre for Disease Prevention and Control also suggested analysis of wastewater from incoming flights as a complement to other monitoring tools to detect introductions of the Omicron variant for the EU/EEA (ECDC, 2021).

## Methods

### Sampling, viral concentration, and DNA extraction

Wastewater samples were collected twice weekly since 30 May 2022 at the wastewater treatment plant (WTP) of Italy’s major airport in Rome (https://www.adr.it/web/aeroporti-di-roma-en/water). The global capacity of this WTP is 4000 m^3^ per day.

In ten weeks, 20 samples (500 ml each) were collected. Twenty-four hours of composite samples were obtained using a refrigerated autosampler. Samples were stored at 4 °C until analysis and processed within three days from sampling. Before viral concentration, samples were pre-treated at 56 °C in a water bath for 30 min to guarantee the safety of the operators. A PEG/NaCl precipitation protocol, originally developed for the environmental surveillance of SARS-CoV-2 (Wu et al., 2020; Cutrupi et al., 2022) was used in this study, with few modifications, consisting in increasing the initial volume of wastewater to 90 ml (2 tubes of 45 ml) and eluting all the extracted nucleic acids in 50 µl. Briefly, samples were centrifuged at 4500*×g* for 30 min to remove solids. PEG8000 (4 g) and NaCl (0.9 g) were then added to the supernatant in each tube and mixed for 15 min. After complete PEG/NaCl dissolution, samples were centrifuged again at 12,000*×g* for 2 h. The pellets of the two tubes were resuspended and pooled with 2 mL of lysis buffer containing guanidine thiocyanate (bioMerieux, France) to achieve viral lysis for nucleic acid extraction. Samples in lysis buffer were incubated for 20 min at room temperature, then 50 μL of magnetic silica beads (bioMerieux) were added to the sample and set for 10 min at room temperature to allow the adhesion of nucleic acids to the beads. A semi-automatic extraction platform (Minimag, bioMerieux) was then used for the subsequent washing phases, and at last, nucleic acids were eluted in 50 μL. To efficiently remove contaminants that can inhibit downstream enzymatic reactions, DNA was further purified using the OneStep PCR Inhibitor Removal Kit (Zymo Research, CA, USA).

### Real-time RT-PCR

Three different Real-Time PCR assays were used in the present study: two published in 2004, targeting the N3R and F3L genes (Kulesh et al., 2004), and one developed in 2010 by the United States Centers for Disease Control and Prevention, targeting the G2R region (TNF receptor gene), named generic real-time PCR assay G2R_G (Li et al. 2010). The G2R assay is among those cited by WHO in the Interim guidance for Monkeypox laboratory testing (WHO 2022c). Upon comparison of the published primers and probes with Monkeypox sequences of the current outbreak, the presence of mismatches in primers and/or probes was highlighted for all the assays, specifically: i) one in the reverse primer of the F3L assay, ii) one in the probe of the N3R assay, and iii) two in the G2R_G assay, in the forward and the reverse primers, respectively.

Therefore, novel primer/probes with 100% nt identity with the current Monkeypox virus outbreak sequences were designed and tested compared to the original ones (Table 1). Monkeypox DNA (Monkeypox virus, 2022, Slovenia ex Gran Canaria, Ref-SKU: 005N-04716, batch 06.06.2022), kindly provided by the European Virus Archive Global (EVAg), was used as a control for primers/probes testing (see Supplementary Materials). After selecting the most efficient primers/probes sets (PCR IDs 1003, 1004, and 1005, Table 1), the assays were further optimized by evaluating different real-time PCR reagents and primer/probe concentrations. Annealing temperatures were set up as in the reference papers (Kulesh et al., 2004; Li et al., 2010). Details of the optimization tests and results are reported in Supplementary Materials.

**Table 1.**
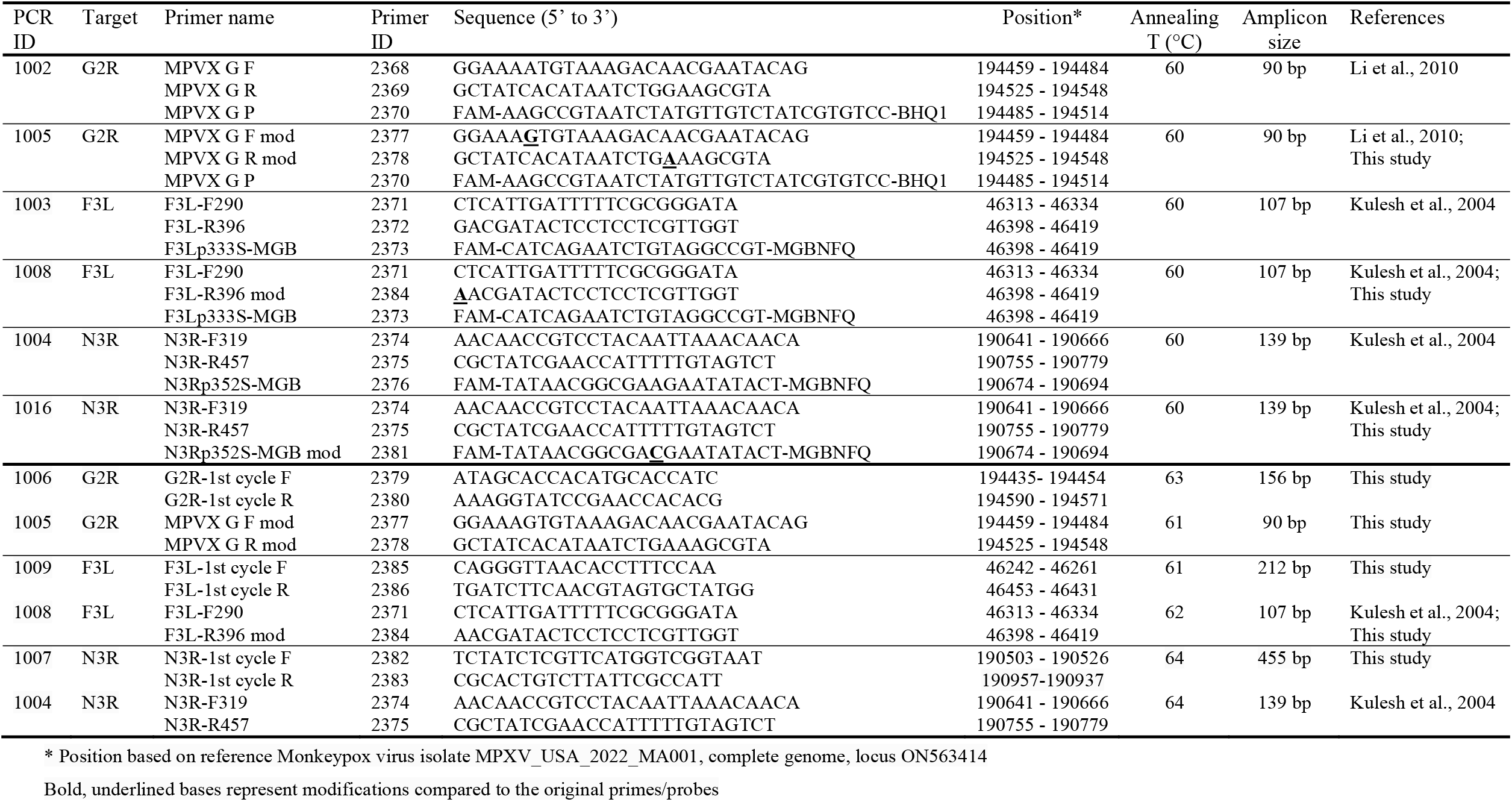
Primers and probes used in the present study.

For environmental samples analysis, reaction mixes were prepared, for all assays, in 25 μL volume using the TaqPath BactoPure Microbial Detection Master Mix (Thermo Fisher Scientific, MA, USA), 800 mM of each primer, 500 nM of the probe and 5 μL of sample DNA. Amplification conditions included an initial activation step at 95 °C for 2 min and 50 cycles of 10 s at 95 °C and 30 s at 60 °C. All real-time assays were run on a QuantStudio 12K Flex (Applied Biosystems, MA, USA). Negative PCR and negative environmental controls were included in each run to monitor potential contamination events. Tenfold dilutions of the standardized EVAg Monkeypox DNA (range 740 – 0.74 g.c/μl) were included in the runs as positive controls and for the rough estimation of Monkeypox viral load in the samples.

For each assay, the limit of detection (LOD_50_) was assessed on pure target (quantified Monkeypox DNA provided by EVAg) and on Monkeypox DNA diluted in nucleic acids previously extracted from wastewater samples collected before the emergence of Monkeypox virus in Europe. Analysis was carried out on serial dilutions, with 8 replicates per dilution (see Supplementary Materials). Calculations of LOD_50_ were done according to Wilrich and Wilrich (2009).

### Nested RT-PCR and sequencing

Three different nested PCR assays targeting the same three regions of the real-time PCR assays were designed to confirm results by amplicon sequencing. Forward and reverse primers used for the real-time PCR assays were used as nested primers. Novel primers were designed upstream and downstream of the amplicon generated by these primers (see Table 1) using the Primer3Plus software (Untergasser et al., 2007), to be used in the first PCR cycle.

Each of the three PCR reactions was performed using 1 μL of 10 μM primers and 2 μL of DNA in a final volume of 25 μL, using the Platinum SuperFi II Green PCR Master Mix (Thermo Fisher Scientific). The amplification conditions for the first PCR (G2R: PCR ID 1006, F3L: ID 1009, and N3R: ID 1007, see Table 1) were as follows: denaturation at 98 °C for 2 min, followed by 39 cycles of 98 °C for 10 s, annealing for 10 s (see temperatures in Table 1), 72 °C for 30 s, and a final extension at 72°C for 5 min. Nested PCR was performed using 2 μL of the first PCR product as template and 1 μL of each 10 μM primer. The following conditions were used for nested PCR amplifications: initial denaturation at 98°C for 30 s, followed by 35 cycles of 98 °C for 10 s, annealing (Table 1) for 30 s, 72 °C for 30 s, and a final extension at 72 °C for 10 min. Standard precautions were taken to avoid laboratory contamination with other templates and amplicons. As an additional precaution to prevent false positive results, no positive control was included during the runs. PCR products were examined by gel electrophoresis (2% agarose gel, stained with GelRed, Biotium; Fremont, CA, USA). Amplicons of the expected length were purified from the PCR reaction using the Montage PCRm96 Micro-well Filter Plate (Millipore, Burlington, MA, USA) and sequenced on both strands (Bio-Fab Research, Rome, Italy, and Eurofins Genomics, Ebersberg, Germany). Forward and reverse sequences were assembled into consensus sequences using Molecular Evolutionary Genetics Analysis version 10 (Kumar et al., 2018), which were classified by BLAST searches.

## Results

All real-time PCR assays successfully amplified the Monkeypox DNA provided by EVAg. The modified primers targeting the F3L and N3R genes did not improve the assay sensitivity (Table 2). Conversely, the modified assay G2R_G, targeting the TNF receptor gene, showed a decrease in the average *Cq* of 1.34 cycles (21.93 *vs*. 23.28) compared to the original assay, demonstrating a better performance. Therefore, subsequent optimization activities and screening of wastewater samples were undertaken using the F3L (PCR ID 1003) and N3R (ID 1004) assays as originally designed (Kulesh et al., 2004), and the G2R_G (ID 1005) assay as modified in the present study.

**Table 2.** Real-time PCR primers comparison.

| PCR ID | Target | Primer name | Primer ID | Average C<sub>q</sub><sup>a</sup> | SD |
| --- | --- | --- | --- | --- | --- |
| 1002 | G2R | MPVX G F | 2368 | 23.28<sup>b</sup> | 0.55 |
|  |  | MPVX G R | 2369 |  |  |
|  |  | MPVX G P | 2370 |  |  |
| 1005 | G2R | MPVX G F mod | 2377 | 21.93 | 0.25 |
|  |  | MPVX G R mod | 2378 |  |  |
|  |  | MPVX G P | 2370 |  |  |
| 1003 | F3L | F3L-F290 | 2371 | 22.95 | 0.31 |
|  |  | F3L-R396 | 2372 |  |  |
|  |  | F3Lp333S-MGB | 2373 |  |  |
| 1008 | F3L | F3L-F290 | 2371 | 23.56 | 0.25 |
|  |  | F3L-R396 mod | 2384 |  |  |
|  |  | F3Lp333S-MGB | 2373 |  |  |
| 1004 | N3R | N3R-F319 | 2374 | 21.76 | 0.16 |
|  |  | N3R-R457 | 2375 |  |  |
|  |  | N3Rp352S-MGB | 2376 |  |  |
| 1016 | N3R | N3R-F319 | 2374 | 26.69<sup>b</sup> | 0.43 |
|  |  | N3R-R457 | 2375 |  |  |
|  |  | N3Rp352S-MGB mod | 2381 |  |  |

<sup>a</sup> Triplicate reactions<sup>b</sup> Duplicate reactions

On pure Monkeypox virus DNA, the real-time PCRs ID 1003, 1004, and 1005 provided a LOD_50_ of 0.21, 0.31 and 0.21 g.c./μl respectively, and of 0.43, 0.33 and 0.31 g.c./μl (2.16, 1.65 and 1.55 g.c./reaction) when testing nucleic acids extracted from sewage samples spiked with the Monkeypox virus.

Results of the real-time PCR assays on the airport wastewaters are reported in Table 3. Two of the 20 wastewater samples tested positive by real-time PCR: sample ID 4445 (collected on June 15) with both the G2R and F3L assays, while sample ID 4478 (collected on July 18) with the G2R assay only. *Cq* values ranged between 38.37 and 40.18, indicating relatively low DNA concentrations in the tested samples (estimated concentrations ≈0.5 and 1.7 g.c./μl of nucleic acid for samples 4445 and 4478, respectively, equivalent to approx. 277 and 944 g.c./L of wastewater).

**Table 3.**
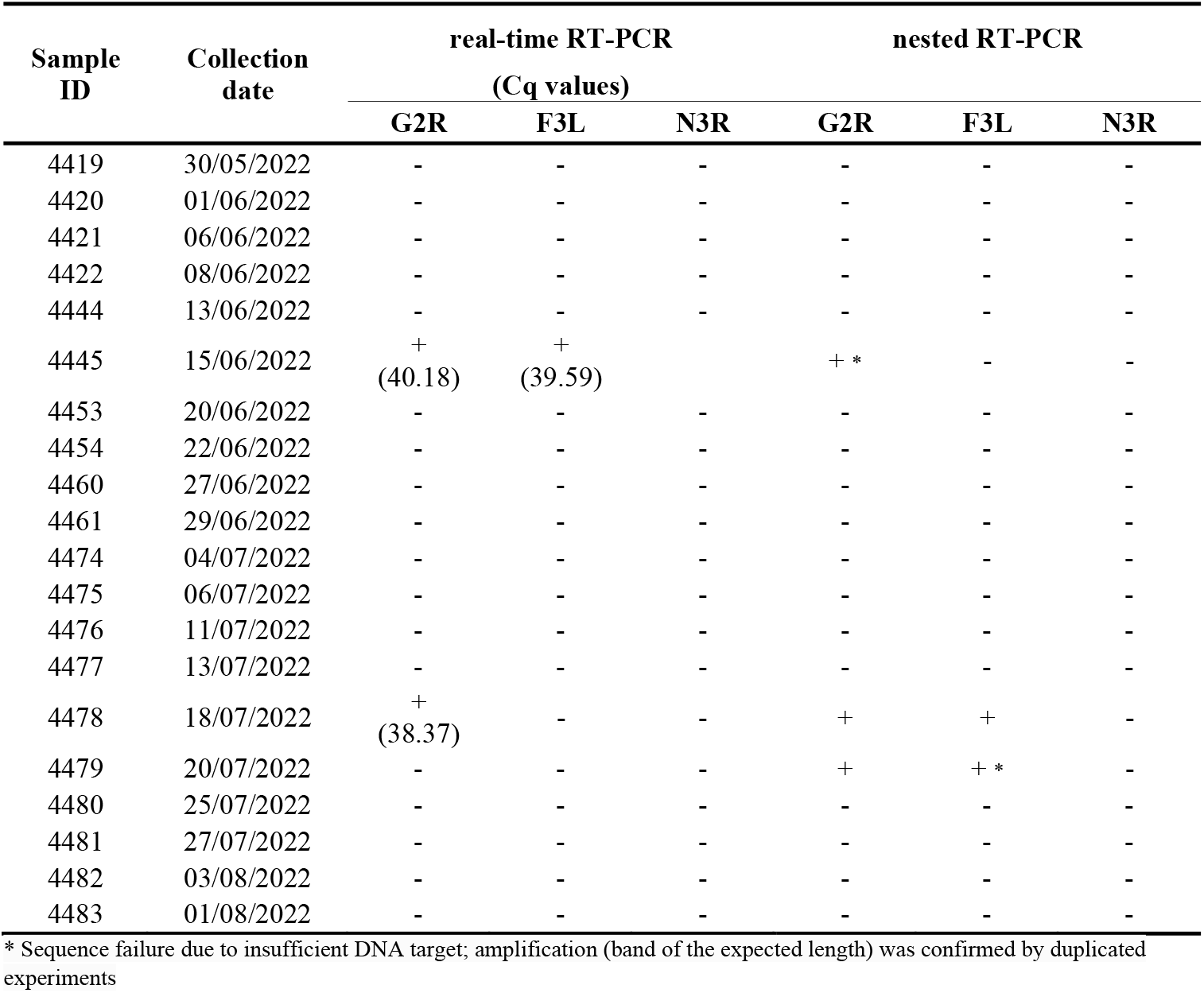
Wastewater sample results.

Amplicons of the expected size were obtained by nested PCR for samples ID 4478 and 4479 (the latter undetected by real-time PCR) with both the F3L and G2R assays, and sample ID 4445 with the G2R assay only. None of the samples could be successfully amplified by the N3R assay. Positive PCR amplicons were subjected to sanger sequencing, and consensus sequences found 100% similarity by BLAST analysis between study sequences and the Monkeypox strains available in GenBank, thus confirming the presence of MPXV DNA.

## Discussion

Monkeypox is a zoonotic disease caused by the MPXV, a double-stranded DNA virus classified in the Orthopoxvirus genus of the Poxviridae family. MPXV was first detected in an outbreak of a vesicular disease among captive monkeys in 1958 (Parker and Buller, 2013). The first human case of Monkeypox was identified in August 1970 in a nine-year-old child with smallpox-like vesicular skin lesions in Zaire (WHO 2022a). Since May 2022, an increasing number of cases of Monkeypox have been reported, affecting non-endemic regions across the globe. Phylogenomic analysis of available MPXV genomes, performed to determine viral evolution and diversity, showed genomes from the ongoing 2022 multi-country outbreak clustering in a newly emerging clade harbouring (Luna et al., 2022). Indeed, we are now facing a multi-country outbreak, with about 18000 cases reported from 41 countries in the European region as of 9 August 2022 (ECDC-WHO, 2022). The Monkeypox Global map of the CDC, updated to the same date, reports 31800 total cases (98.8% of which are in countries with no history of monkeypox cases) and 89 countries involved, 82 of which reported the first occurrence of the virus (CDC 2022). The World Health Organization declared Monkeypox a public health emergency of international concern, suggesting a more coordinated global response, further research, and the ramping up of vaccine production (WHO 2022b). A crucial aim of infectious disease surveillance is the early detection of cases, which is essential for disease control. Rapid detection, isolation, testing, and management of cases are needed for a timely response. In the present study, we aimed to provide insights into the possibility of monitoring MPXV through wastewater surveillance, a well-established complementary epidemiological tool for other viral infectious diseases such as SARS-CoV-2 (Bibbi et al., 2021; Bonanno Ferraro et al., 2021; Dutta et al., 2021; Lundi et al., 2020; Medema et al., 2020; Shah et al., 2021). The rationale behind wastewater surveillance is based on the observation that individuals with an infection, symptomatic or not, may shed viruses in feces or other bodily fluids that, through sewer systems, end up in WTPs. It is, therefore, possible to monitor virus circulation in the population by examining sewage samples. Individuals infected with Monkeypox excrete the virus via skin lesions, feces, urine, and saliva (Peiró-Mestres et al., 2022; Lapa et al., 2022; Antinori et al., 2022), suggesting that MPXV is likely to be detected in wastewater. We focused our study on investigating wastewater samples obtained from a airline transportation hub, where many people from different countries transit. Since 30 May 2022, wastewater samples collected from a biological treatment plant located in Rome, serving Italy’s first airport, were tested for MPXV. Three of the 20 samples tested positive for MPXV by real-time (low viral concentrations) and/or nested PCR followed by sequencing. Harmonized methods for detecting MPXV in wastewater are not yet available. Therefore, we tested three different real-time PCR assays previously designed for clinical samples (Li et al., 2020; Kulesh et al., 2004). Before testing environmental samples, we modified all assays by introducing small changes in the primer/probe sequences to mitigate the effect of nucleotide mismatches. MPXV DNA provided by the EVAg project was used as a positive control. While no increase in assay sensitivity was detected for the modified N3R and F3L assays (Kulesh et al., 2004), the modified MPXV generic G2R_G assay (Li et al., 2010), targeting the TFN gene, provided an improvement in the detection signal. This assay (as originally designed) had already demonstrated higher sensitivity than other real-time PCR assays, possibly in relation to the fact that two copies of this gene are present in the MPXV genome due to its location at the ITR terminal regions (Li et al., 2010). Therefore, the optimized G2R_G assay and the original N3R and F3L assays (Kulesh et al., 2004) were used to test environmental samples. The usefulness of monitoring transportation hubs has already been demonstrated for SARS-CoV-2 in airports or ships (Ahmed et al., 2020; Agrawal et al., 2022), and the European Centre for Disease Prevention and Control also suggested analysis of wastewater from incoming flights to detect introductions of the SARS-CoV-2 Omicron for the EU/EEA (ECDC, 2021). We confirm monitoring transportation hubs a powerful tool for monitoring infections traveling from one location to another. Indeed, public transportation systems link all regions on Earth, making it possible for people to travel around the world to serve as a network through which an epidemic can spread worldwide (Xu et al., 2013).

Although fragments of Monkeypox virus DNA were found in wastewater in the present study, this does not imply the risk of transmission through the wastewater. Indeed, the standard treatments and disinfectant processes at WTPs are effective for the inactivation of viruses. Moreover, Monkeypox virus is not known to have a waterborne transmission route, and according to ECDC it does not spread easily between people: human-to-human transmission occurs through close contact with infectious material from skin lesions of an infected person, through respiratory droplets in prolonged face-to-face contact, and through fomites (ECDC 2022).

In Italy, the first monkeypox case was confirmed on 20 May 2022. Early on, the Ministry of Health activated a monkeypox surveillance system with the collaboration of Regions and Autonomous Provinces, publishing a bulletin on confirmed cases every Tuesday and Friday (Vaiolo delle scimmie (salute.gov.it). As of 9 August 2022, 599 cases have been confirmed in Italy (with a 94 unit increase compared to the ECDC-WHO bulletin published on 2 August), 169 of which related to subjects traveling abroad. Cases have been documented in 17 of the 21 Italian Regions/Autonomous provinces. However, asymptomatic or paucisymptomatic infections may go undetected; moreover, individuals with clinical manifestations may decide not to have healthcare consultation; therefore, some infections are not reported, and cases are underestimated. Environmental surveillance of Monkeypox applied on a national scale could help track Monkeypox infections and sound the alarm for the introduction of the virus in a new region or the increase in the number of cases.

To our knowledge, no peer-reviewed articles have been published on detecting MPXV DNA in wastewater as of 12 August 2022. However, two preprints, submitted in July 2022, are available. Wolfe and co-workers quantified Monkeypox virus DNA in settled solids samples from nine wastewater plants in California over four weeks (Wolfe et al., 2022). They detected MPXV DNA at nearly all the wastewater plants investigated. They used the G2R_G assay developed by the CDC (Li et al., 2010) that we optimized in the present study. In Europe, de Jonge and collaborators used the same G2R assay to detect MPXV DNA in urban wastewater samples collected in the Netherlands and, occasionally, from the Amsterdam Schiphol Airport, a major travel hub (de Jonge et al., 2022). Furthermore, the Indian COVID-19 Technical Advisory Committee has recommended wastewater surveillance for early detection of Monkeypox, suggesting beginning with samples from the International Airport, the entry point of international arrivals (TAC recommends sewage surveillance for early detection of Monkeypox in State - The Hindu). Researchers in Thailand are also examining wastewater for signs of Monkeypox (Thai researchers test wastewater to track spread of monkeypox| Monkeypox | The Guardian).

In conclusion, our data show that detecting MPXV DNA in wastewater is feasible. Increasing levels of monkeypox in wastewater will alert us, providing time for health systems to organize and react. Results confirm that environmental monitoring through transportation hubs has the potential to track the introduction and/or the diffusion of emerging biological hazards. The next stage will be testing wastewater samples originating from WTPs distributed throughout Italy to monitor MPXV geographical spread in the country. In the future, studies on the occurrence of MPXV in wastewater are likely to increase. Next efforts should focus on understanding how the detection of the viral DNA in sewage can be related to reported/confirmed cases. Results suggest that wastewater surveillance can be quickly adapted to investigate new viral pathogens.

## Data Availability

All data produced in the present study are available upon reasonable request to the authors

## Acknowledgments

This publication was supported by the European Virus Archive Global (EVA-GLOBAL) project, which provided the Monkeypox DNA for testing the Real-time PCR assays. For wastewater sample collection, we gratefully acknowledge the support of Cavina Lorenzo and Eleuteri Silvia, Aeroporti di Roma (ADR) Fiumicino Airport. We also thank Claudia Del Giudice, Lidia Orlandi and Serena Maccaroni for their technical assistance. This research was partially supported by collaboration agreement EC G.A. NO. 060701/2021/864481/ SUB/ ENV.C2 - “Support to Member States for the creation of systems, local collection points and digital infrastructures for monitoring COVID 19 and its variants in wastewater - Italy”.

## SUPPLEMENTARY MATERIALS

### 1) Comparison of primers/probes sets

To select the primers/probes sets providing better real-time PCR results, a 10^−2^ dilution of the Monkeypox DNA Slovenia ex Gran Canaria, Ref-SKU: 005N-04716 provided by EVAg was tested as control, using PCR IDs 1002, 1003, 1004, 1005, 1008, 1016 (see Table 1 in the main text).

For the sake of comparison, all primers/probes sets were assayed in the same conditions: real-time reactions were prepared in 25 μL volume using the TaqPath BactoPure Microbial Detection Master Mix (Thermo Fisher Scientific, MA, USA), 500 mM of each primer, 250 nM of probe and 5 μL of sample DNA. Amplification conditions included an initial activation at 95 °C for 2 min, and 45 cycles of 10 s at 95 °C and 30 s at 60 °C. Each reaction was run in triplicate on a QuantStudio 12K Flex (Applied Biosystems, MA, USA).

The three primers/probes sets with the lowest *Cq* value were selected for further optimization: PCR IDs 1003 (F3L), 1004 (N3R) and 1005 (G2R).

### 2) Standardization of Monkeypox DNA provided by the European Virus Archive Global (EVAg)

To determine the Limit of Detection (LOD_50_) of the selected real-time PCR, the Monkeypox DNA (Monkeypox virus, 2022, Slovenia ex Gran Canaria, Ref. 005V-04714 batch 06.06.2022) provided by EVAg was quantified by means of droplet digital PCR (ddPCR).

Tenfold dilutions of the DNA stock were prepared in molecular grade Tris-EDTA buffer (10 mM Tris, 1 mM EDTA) pH 8.0 (Sigma-Aldrich).

DNA dilution 10^−2^, 10^−3^, 10^−4^ and 10^−5^ were tested in quadruplicate by ddPCR performed using the BioRad QX200 system (Bio-rad, CA, USA) and the ddPCR Supermix for probes kit – no dUTP (BioRad). The reaction mixture included: 10 μl ddPCR supermix, primers 500 nM, probes 250 nM and nuclease-free water to a final volume of 20 μl. Primers and probes were those described for PCR IDs 1003, 1004 and 1005.Droplets were generated as recommended by the manufacturer and amplification was performed on a 9600 GenAmp thermalcycler (Applied BioSystems) as follows: 95 °C for 10 min followed by 94 °C for 30 sec and 60 °C for 60 sec (40 cycles) and by a final stage at 98 °C for 10 min. Results were acquired using the Bio-Rad QX200

Droplet Reader and QX Mangaer Standard Edition (v1.2) to provide absolute quantification of the target sequence.

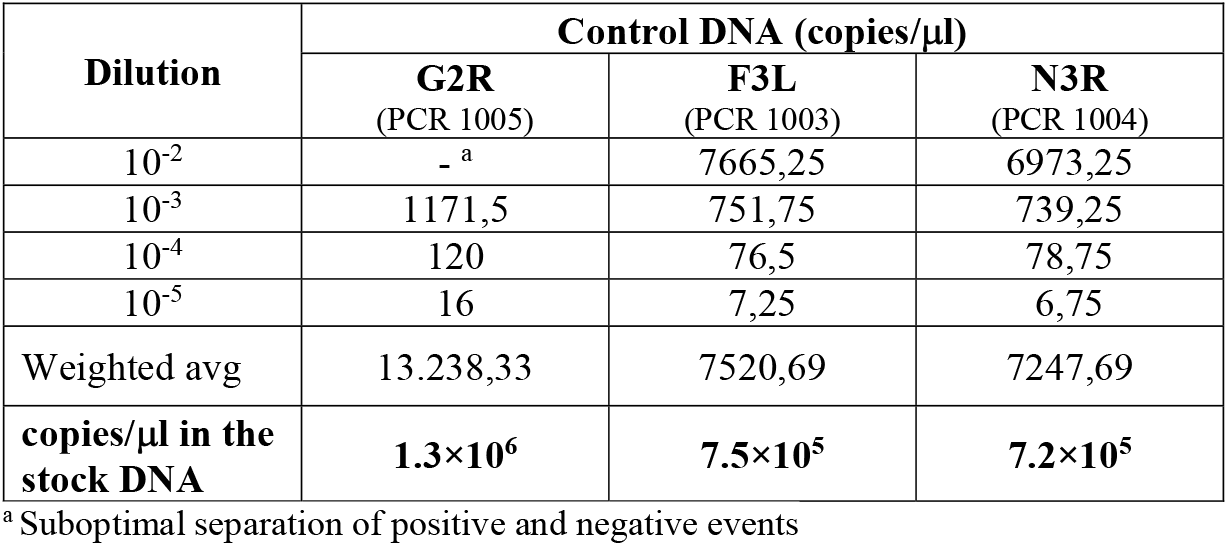

| Dilution | Control DNA (copies/μl) |  |  |
| --- | --- | --- | --- |
|  | G2R<br>(PCR 1005) | F3L<br>(PCR 1003) | N3R<br>(PCR 1004) |
| 10<sup>-2</sup> | - <sup>a</sup> | 7665,25 | 6973,25 |
| 10<sup>-3</sup> | 1171,5 | 751,75 | 739,25 |
| 10<sup>-4</sup> | 120 | 76,5 | 78,75 |
| 10<sup>-5</sup> | 16 | 7,25 | 6,75 |
| Weighted avg | 13.238,33 | 7520,69 | 7247,69 |
| **copies/μl in the stock DNA** | **1.3×10<sup>6</sup>** | **7.5×10<sup>5</sup>** | **7.2×10<sup>5</sup>** |

<sup>a</sup> Suboptimal separation of positive and negative events

Since two copies of the G2R region are present in Monkeypox virus genome due to its location at the ITR terminal regions, only the results of F3L and N3R were taken into account for the quantification of viral genome copies (g.c.) of the Monkeypox Slovenia ex Gran Canaria, Ref-SKU: 005N-04716 provided by EVAg. Reference value was defined as the average of the values obtained for the two targets: 738419 g.c./μL = **7.4×10**^**5**^ **g.c./μl**

### 3) Optimization of real-time PCR ID 1003 (F3L), 1004 (N3R) and 1005 (G2R) conditions

#### a) Testing of different real-time reagents

Based on previous experiences, the choice of real-time reagents may affect the efficiency of target virus detection in wastewater samples, due to different sensitivity of polymerases to factors as environmental inhibitors or supercoiling of target sequences. To select the most efficient reagents for Monkeypox DNA detection in wastewater, the standardized EVAg Monkeypox DNA was diluted in a 1:100 proportion in nucleic acids extracted from wastewater samples collected from urban WTPs in a period preceding the emergence of Monkeypox virus in Italy (November and December 2021).

Samples were then tested with PCR IDs 1003, 1004 and 1005 using the following reagents:

– AgPath-ID One-Step RT-PCR Reagents (Applied Biosystems, MA, USA)
– TaqPath BactoPure Microbial Detection Master Mix (Thermo Fisher Scientific, MA, USA)
– TaqMan Fast Advanced Master Mix (Thermo Fisher Scientific, MA, USA)

All reactions were prepared in 25 μL volume using 500 mM of each primer, 250 nM of probe and 5 μL of sample DNA. AgPath-ID reactions included 1 μl/rxn of enzyme mix and 1.67 μl of detection enhancer. Reactions were run on a QuantStudio 12K Flex (Applied Biosystems, MA, USA). Amplification conditions included:

– AgPath-ID One-Step RT-PCR Reagents: an initial reverse transcriptase inactivation step at 95°C for 5 min, followed by 45 cycles of 10 s at 95 °C and 30 s at 60 °C.
– TaqPath BactoPure Microbial Detection Master Mix: an initial activation step at 95 °C for 2 min, followed by 45 cycles of 10 s at 95 °C and 30 s at 60 °C.
– TaqMan Fast Advanced Master Mix: an initial hold at 50 °C for min, followed by an activation step at 95 °C for 20 s, and by 45 cycles of 10 s at 95 °C and 30 s at 60 °C.

The following Cq values were obtained:

| Reagent | G2R<br>(PCR 1005) | F3L<br>(PCR 1003) | N3R<br>(PCR 1004) |
| --- | --- | --- | --- |
| AgPath-ID One-Step RT-PCR | 21.71 | 23.07 | 21.60 |
| TaqPath BactoPure Microbial Detection Master Mix | 21.90 | 23.18 | 21.91 |
| TaqMan Fast Advanced Master Mix | 22.20 | 22.60 | 21.78 |

As no significant difference was detected among the performance of the different reagents, TaqPath BactoPure Microbial Detection Master Mix was selected, as it required the least preparation time.

#### b) Primers/probes concentration

Optimization of primers/probes concentration was performed by testing, for each PCR assay (IDs 1003, 1004 and 1005), the following concentrations in their different combinations:

– primer forward: 200 nM, 500 nM, 800 nM
– primer reverse: 200 nM, 500 nM, 800 nM
– probe: 100 nM, 250 nM, 500 nM

All reactions were prepared in 25 μL volume using 5 μL of sample DNA (10^−4^ dilution of the standardized EVAg Monkeypox DNA) and the TaqPath BactoPure Microbial Detection Master Mix. Amplification conditions included an initial activation step at 95 °C for 2 min, followed by 45 cycles of 10 s at 95 °C and 30 s at 60 °C. Reactions were run on a QuantStudio 12K Flex (Applied Biosystems, MA, USA).

Results of the tests are graphically summarized in Figure A (F3L, PCR ID 1003), B (N3R, PCR ID 1004) and C (G2R, PCR ID 1005):

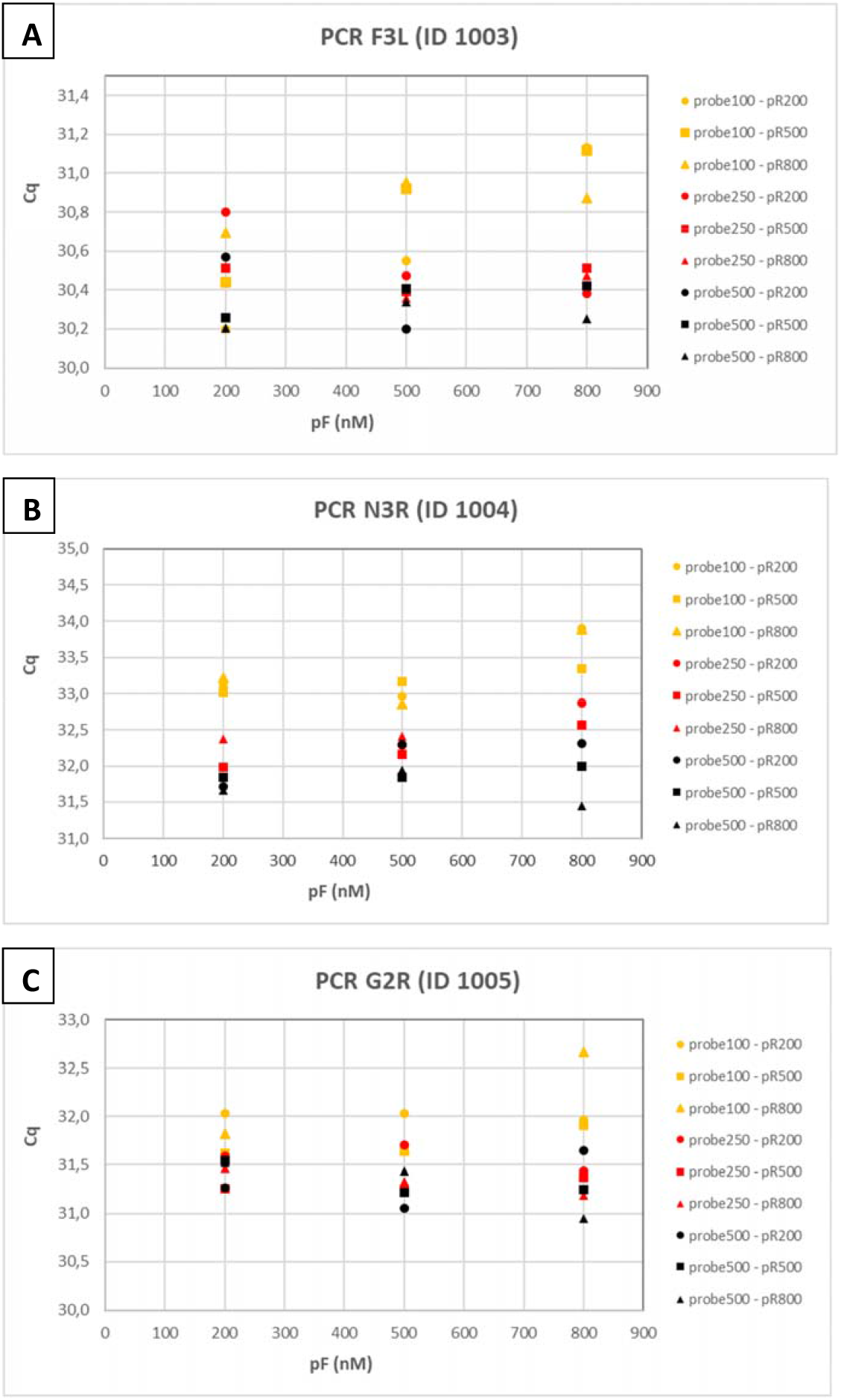

Based on *Cq* values, in all PCR assays, better amplifications were achieved at a probe concentration of 500 nM and slightly better results were obtained with a primer concentration of 800 nM although, in the latter case, the differences with other concentrations were minimal (often below 1 ΔCq).

Therefore, concentrations of 500 nM of probe and 800 nM of primers were used for the analysis of the environmental samples.

### 4) Assessment of LOD50 for real-time PCR ID 1003 (F3L), 1004 (N3R) and 1005 (G2R)

#### a) LOD_50_ on pure target (Monkeypox DNA diluted in TE buffer)

To assess the sensitivity of the real-time PCR assays used in the study, the LOD_50_ of each reaction was calculated according to Wilrich and Wilrich (2009).

Two-fold dilutions of the standardized EVAg Monkeypox DNA in molecular grade TE buffer pH 8.0 were prepared starting from the 10^−5^ dilution (7.4 g.c./μl). Each dilution was tested in 8 replicates with PCR ID 1003 (F3L), 1004 (N3R) and 1005 (G2R), using the optimized reaction conditions.

The following results were obtained:

| Expected target concentration (g.c./μl) | 1005 (G2R) | 1003 (F3L) | 1004 (N3R) |
| --- | --- | --- | --- |
| 3,7 | 8 / 8 | 8 / 8 | 8 / 8 |
| 1,85 | 8 / 8 | 8 / 8 | 8 / 8 |
| 0,925 | 7 / 8 | 8 / 8 | 8 / 8 |
| 0,4625 | 7 / 8 | 5 / 8 | 7 / 8 |
| 0,23125 | 4 / 8 | 2 / 8 | 3 / 8 |
| 0,115625 | 3 / 8 | 1 / 8 | 2 / 8 |
| **LOD<sub>50</sub> (g.c./μl)** | **0.213** | **0.306** | **0.206** |
| **LOD<sub>95</sub> (g.c./μl)** | **0.923** | **1.323** | **0.890** |
| **LOD<sub>50</sub> (g.c./rxn)** | **1.065** | **1.530** | **1.030** |
| **LOD<sub>95</sub> (g.c./rxn)** | **4.615** | **6.615** | **4.450** |

The LOD_50_ of each reaction was calculated using the spreadsheet available at https://www.wiwiss.fu-berlin.de/fachbereich/vwl/iso/ehemalige/wilrich/index.html.

#### b) LOD_50_ on target in wastewater (Monkeypox DNA diluted in nucleic acids extracted from wastewater samples)

To assess the sensitivity of the real-time PCR assays in the condition of use (i.e. detecting Monkeypox virus in wastewater samples), the LOD_50_ of each reaction was also calculated – with the same approach previously described – by testing the target Monkeypox DNA diluted in nucleic acids extracted from wastewater samples, which include potential inhibitors of the polymerization reaction.

Two-fold dilutions of the standardized EVAg Monkeypox DNA in molecular grade TE buffer pH 8.0 were prepared starting from the 10^−4^ dilution (74 g.c./μl). Each dilution was then used to spike in 1:10 proportion nucleic acids extracted from wastewater samples collected from urban WTPs in a period preceding the emergence of Monkeypox virus in Italy (November and December 2021). Spiked samples were tested in 8 replicates with PCR ID 1003 (F3L), 1004 (N3R) and 1005 (G2R), using the optimized reaction conditions.

The following results were obtained:

| Expected target concentration (g.c./μl) | 1005 (G2R) | 1003 (F3L) | 1004 (N3R) |
| --- | --- | --- | --- |
| 7,4 | 8 / 8 | 8 / 8 | 8 / 8 |
| 3,7 | 8 / 8 | 8 / 8 | 8 / 8 |
| 1,85 | 8 / 8 | 8 / 8 | 8 / 8 |
| 0,925 | 6 / 8 | 7 / 8 | 7 / 8 |
| 0,4625 | 3 / 8 | 6 / 8 | 5 / 8 |
| 0,23125 | 3 / 8 | 1 / 8 | 3 / 8 |
| **LOD50 (g.c./μl)** | **0.432** | **0.330** | **0.309** |
| **LOD95 (g.c./μl)** | **1.868** | **1.427** | **1.336** |
| **LOD50 (g.c./rxn)** | **2.160** | **1.650** | **1.545** |
| **LOD95 (g.c./rxn)** | **9.340** | **7.135** | **6.680** |

Calculations of LOD_50_ were performed as previously reported.

## Notes

### Competing Interest Statement

The authors have declared no competing interest.

